# Association between prescription drugs and all-cause mortality risk in the UK population

**DOI:** 10.1101/2024.03.08.24303967

**Authors:** Jonas Morin, Yves Rolland, Heike A. Bischoff-Ferrari, Alejandro Ocampo, Kevin Perez

**Affiliations:** EPITERNA, Epalinges, Switzerland; IHU HealthAge, UMR CERPOP 1295, CHU Toulouse, Toulouse, France; Department of Geriatric Medicine and Aging Research, University of Zurich, Zurich, Switzerland; Department of Biomedical Sciences, Faculty of Biology and Medicine, University of Lausanne, Lausanne, Switzerland

**Author notes:** Correspondence: A.O., K.P.

## Abstract

Although most drugs currently approved are meant to treat specific diseases or symptoms, it has been hypothesized that some might bear a beneficial effect on lifespan in healthy older individuals, outside of their specific disease indication. Such drugs include, among others, metformin, SGLT2 inhibitors and rapamycin. The UK biobank has recorded prescription medication and mortality data for over 500’000 patients during more than 40 years. In this work we examined the impact of the top 406 prescribed medications on overall mortality rates within the general population of the UK. As expected, most drugs harbor a negative effect on lifespan, probably due to the underlying negative effect of the disease the drug is intended for. Importantly, a few drugs seem to have a beneficial effect on lifespan, including notably Sildenafil, Atorvastatin, Naproxen and Estradiol. These retrospective results warrant further investigation in randomized controlled trials.

## Introduction

With an increasingly aging population, aging poses a significant challenge on society and health care systems^1^, aging being the major risk factor for all age-related diseases. With aging also comes a slew of decline in functional ability (mobility, cognition, and immunity), and a dramatic increase in mortality risk. Promoting healthy aging through primary prevention and lifestyle modifications (avoidance of smoking and alcohol, exercise, diet, sleep) is a well-demonstrated but also complicated strategy to implement. It is then also important to identify drugs that can prevent the functional decline associated with aging, delay the occurrence of age-related diseases, and reduce mortality risk in the elderly. Medications are usually associated with a higher mortality rate. This relation can be explained, in part, by the potential side-effects of the drugs, but mainly because the underlying diseases the drugs are prescribed to treat have themselves an increased mortality risk, that the drug is not able to counteract. Furthermore, increased medication in the population is generally seen as an indicator of poor health, and polypharmacy negatively impacts life expectancy in old people^2,3^.

However, following the “Geroscience hypothesis”^4^, some long-established drugs are being proposed to be tested for their protective effect on aging and mortality in a healthy aging population, including metformin, rapamycin, sodium/glucose cotransporter 2 (SGLT2) inhibitors, acarbose, angiotensin-converting enzyme (ACE) inhibitors, or senolytics^5,6^. Aspirin was notably tested for its beneficial effect on survival in a large, randomized trial in older adults, but with negative results^7^. In a retrospective study, metformin was shown to provide increased overall survival, even compared to non-diabetic control subjects^8^. This finding suggests that other approved drugs could be repurposed to decrease the mortality risk in healthy middle-aged or elderly adults. Yet, one of the main challenges is that drugs are typically prescribed to patients for a specific disease indication, who may already have an increased mortality risk due to this underlying disease, thus confounding the potential beneficial effect on lifespan.

Previous efforts have been made to evaluate the effects of medications on mortality in the general population, but most studies tend to focus on specific classes of medications used for a particular disease^9–12^. Notably, the effect of specific medications on mortality risk has been assessed previously for cancer treatments in cancer patients^13^, statins in patients with or at risk for cardiovascular diseases ^10,14,15^, anti-psychotic and anti-depressant drugs in schizophrenic patients^12,16^, diabetic drugs in diabetic patients^8,11,17,18^. To date, to the best of our knowledge, a study that comprehensively looks at a wide range of commonly prescribed medications and their association with mortality in the general population is missing.

The UK Biobank (UKBB) is a large-scale biomedical database and research resource containing de-identified genetic, lifestyle and health information from half a million UK participants. More precisely, it contains prescription medication data, together with mortality data in the general UK population, in adults aged 37 to 73, for over 500’000 subjects. We used this resource to assess the effect of medication on mortality risk in the general population for 406 of the most prescribed drugs in the UK. Moreover, we also analyze dose-response effects, gender specific effects, and medication class effects.

## Results

We obtained data from the UK Biobank on October 24, 2023, including prescription medication records, mortality data, co-morbidities, and lifestyle factors. The UK Biobank study included 501’169 participants recruited between 37 and 73 years old. Data on 56’213’338 prescriptions were available for 222’058 participants. We excluded participants for whom clinical data was unavailable, as well as accidental or self-inflicted deaths. To identify potential confounders, we first assessed what factors were contributing the most to survival in the general population. 8.4% of patients died during follow-up. The age at recruitment was 57yo. (SD 8yo.). 46% were men, 8% had a cancer diagnosis, 5% had diabetes, 10% were current smokers (**Table S1)**. Not surprisingly, among the factors that were available for analysis (**Methods**), current smoking (HR 2.00, CI 1.94-2.06), cancer diagnosis (HR 1.88, CI 1.83-1.94), diabetes (HR 1.65, CI 1.60-1.70), male gender (HR 1.64, CI 1.60-1.67), and older age at recruitment (HR 1.72, CI 1.69-1.76) were associated with reduced lifespan (**Figure S1, Data Table 1**).

After harmonization (**Methods, Data Table 5**), we found 406 distinct drugs prescribed to more than 500 patients, for a duration of at least 3 months (from the date between first and last use). The 3 most prescribed drugs in the dataset were amoxicillin (N=73371), simvastatin (N=45776), and omeprazole (N=44100) (**Data Table 2**). For each drug we then assessed one by one the survival of patients taking the drug, compared to health-matched controls not taking the drug. The matching was based on covariates identified above to have the strongest impact on survival, namely current smoking, cancer diagnosis, diabetes, gender, age at recruitment (**Methods**). The cohort’s health characteristics varied greatly depending on the different prescription drugs. For example, Atorvastatin patients had a higher age at recruitment (61yo. vs 57yo. for the whole cohort), higher percentage of males (58% vs 46%), and high percentage of diabetes (15% vs 5%). The patient matching strategy was therefore critical to be able to correctly assess the effect of each drug (**Figure S2**).

Out of the 406 drugs studied, 169 had a significant effect on lifespan, after multiple comparison correction (FDR < 0.05). From these 169, as expected, the majority (N = 155, 92%) were associated with increased mortality (**Figure 1, Data Table 2**). These included notably opioids like Morphine sulfate (HR 5.56, CI 4.51-6.86) and Oramorph (HR 5.38, CI 4.08-7.09), the diuretic Furosemide (HR 2.00, CI 1.86-2.15), pain medication Paracetamol (HR 1.48, CI 1.42-1.55) or chronic obstructive pulmonary disease treatment Tiotropium (HR 1.96, CI 1.77-2.17). Importantly, we identified 14 drugs that increased lifespan, compared to health matched controls (**Figure 2, Data Table 2**), independently of current smoking, cancer diagnosis, diabetes, gender, and age at recruitment. These included notably the statin Atorvastatin (HR 0.91, CI 0.87-0.95), the PDE5 inhibitor Sildenafil (HR 0.85, CI 0.78-0.93), the anti-inflammatory drug Naproxen (HR 0.90, CI 0.85-0.96), and the estrogen related drugs Estraderm (HR 0.67, CI 0.51-0.88), Vagifem (HR 0.73, CI 0.59-0.91), Estriol (HR 0.74, CI 0.60-0.92) and Estradiol (HR 0.75, CI 0.59-0.95). Others included, Lymecycline, Otomize, Marvelon, and 2 vaccines (Avaxim, Revaxis).

**Figure 1.**
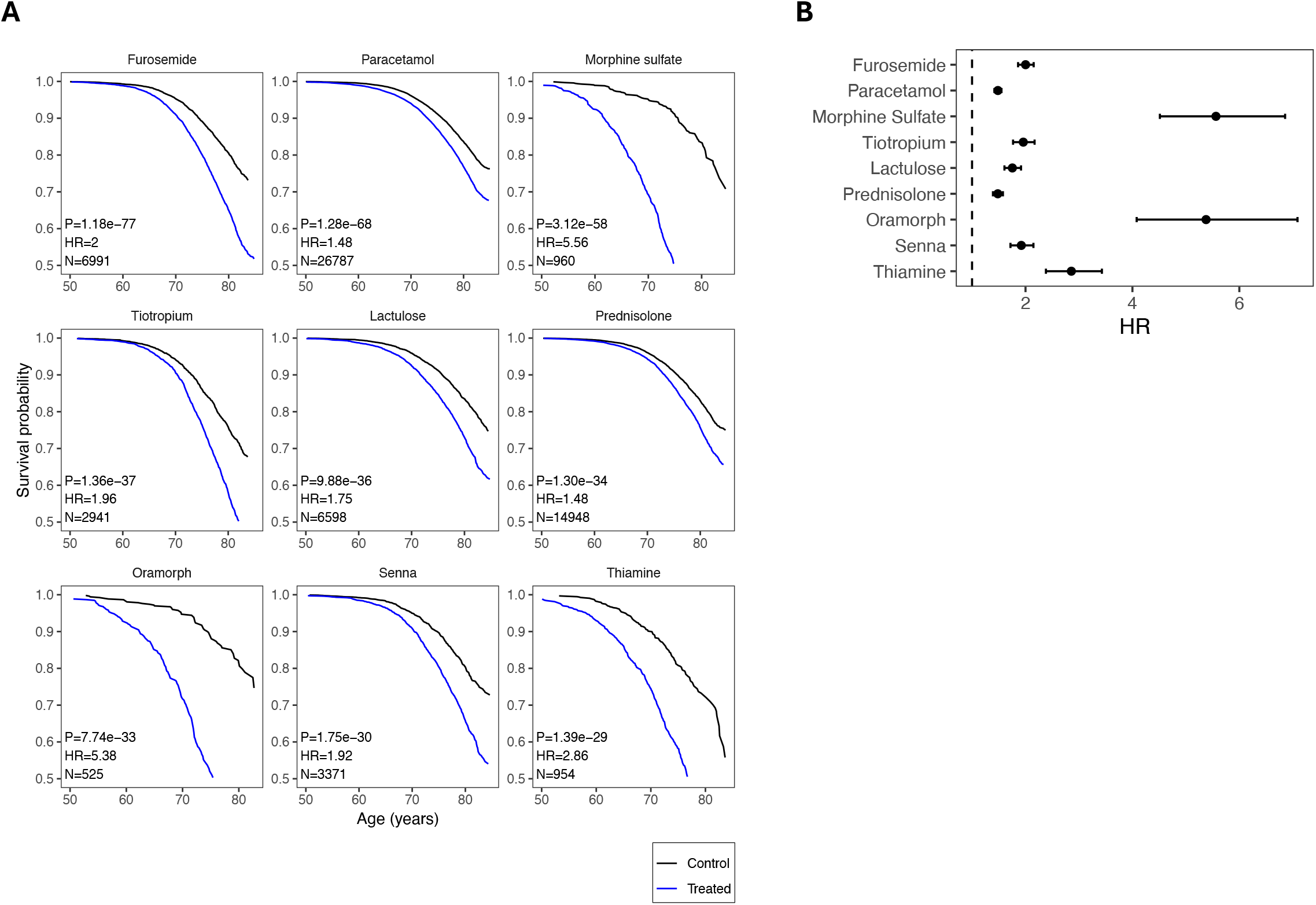
Prescription drugs associated with increased mortality. Top 9 prescription drugs negatively impacting lifespan, ranked by p-value in the Cox Proportional Hazards (CoxPH) model. (**A**) Survival curves. Survival probability vs. Age (years). N: sample size (treated), Hazard-Ratio (HR), > 1 for worse survival, P-value of CoxPH model. Treated group in blue, control in black (**B**) HR with confidence interval 95%. Dotted line HR = 1.

**Figure 2.**
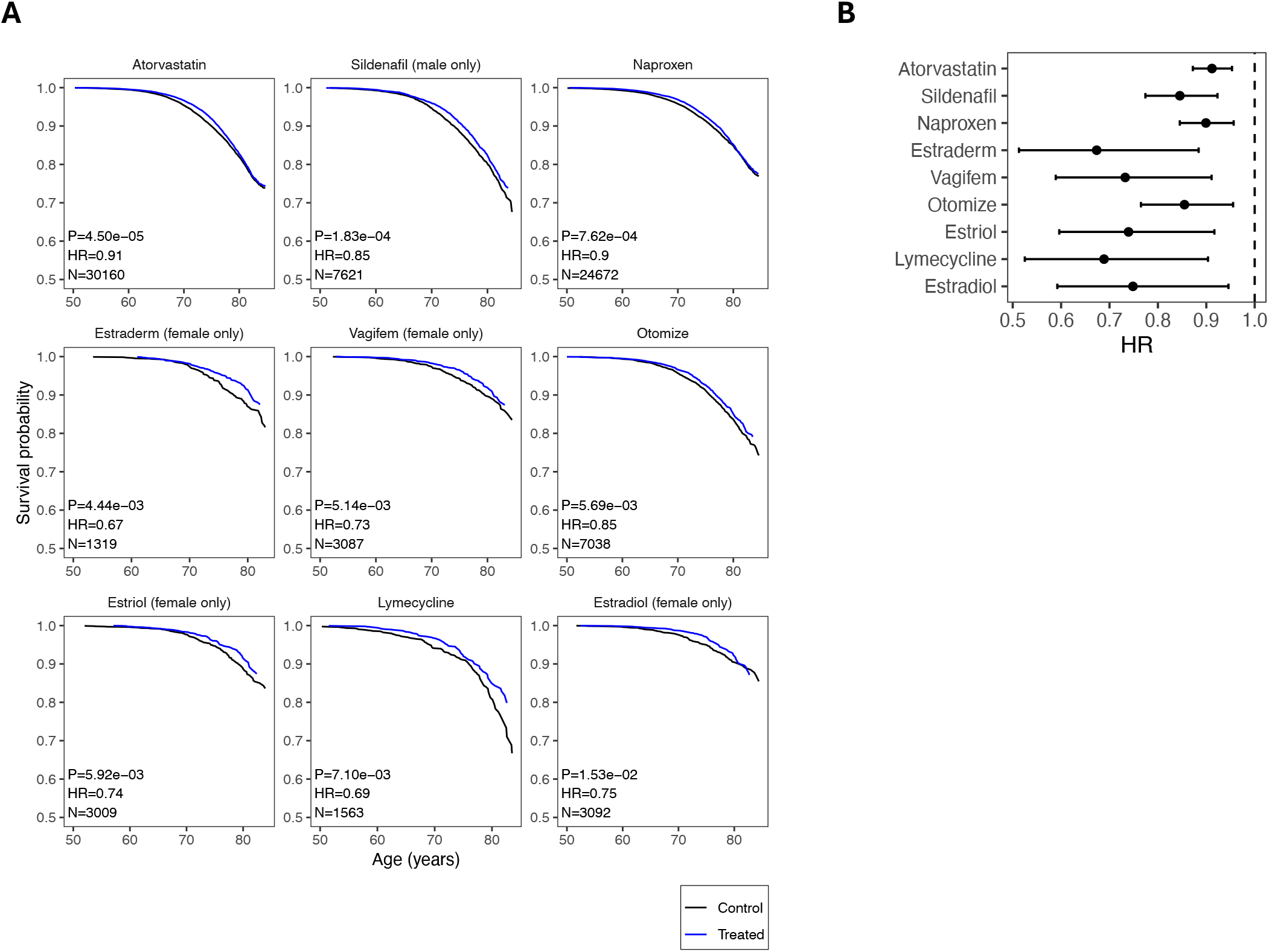
Prescription drugs associated with decreased mortality. Top 9 prescription drugs positively impacting lifespan, ranked by p-value in the Cox Proportional Hazards (CoxPH) model. (**A**) Survival curves. Survival probability vs. Age (years). N: sample size (treated), Hazard-Ratio (HR), > 1 for worse survival, P-value of CoxPH model. Treated group in blue, control in black (**B**) HR with confidence interval 95%. Dotted line HR = 1.

Estrogen related drugs are approved for women only, while PDE5i are for men only, therefore it was possible to look at their effect in one gender only. For drugs approved for both genders, we sought to investigate whether the effect on lifespan was gender-specific or consistent between men and women. Interestingly Atorvastatin increased lifespan, in both males (HR 0.93, CI 0.88-0.98) and females (HR 0.88, CI 0.81-0.95). Conversely for Naproxen the protective effect was more pronounced in men (HR 0.87, CI 0.80-0.95) than in women, and for Otomize more pronounced in women (HR 0.78, CI 0.65-0.93) than in men (**Figure 3, Data Table 2**).

**Figure 3.**
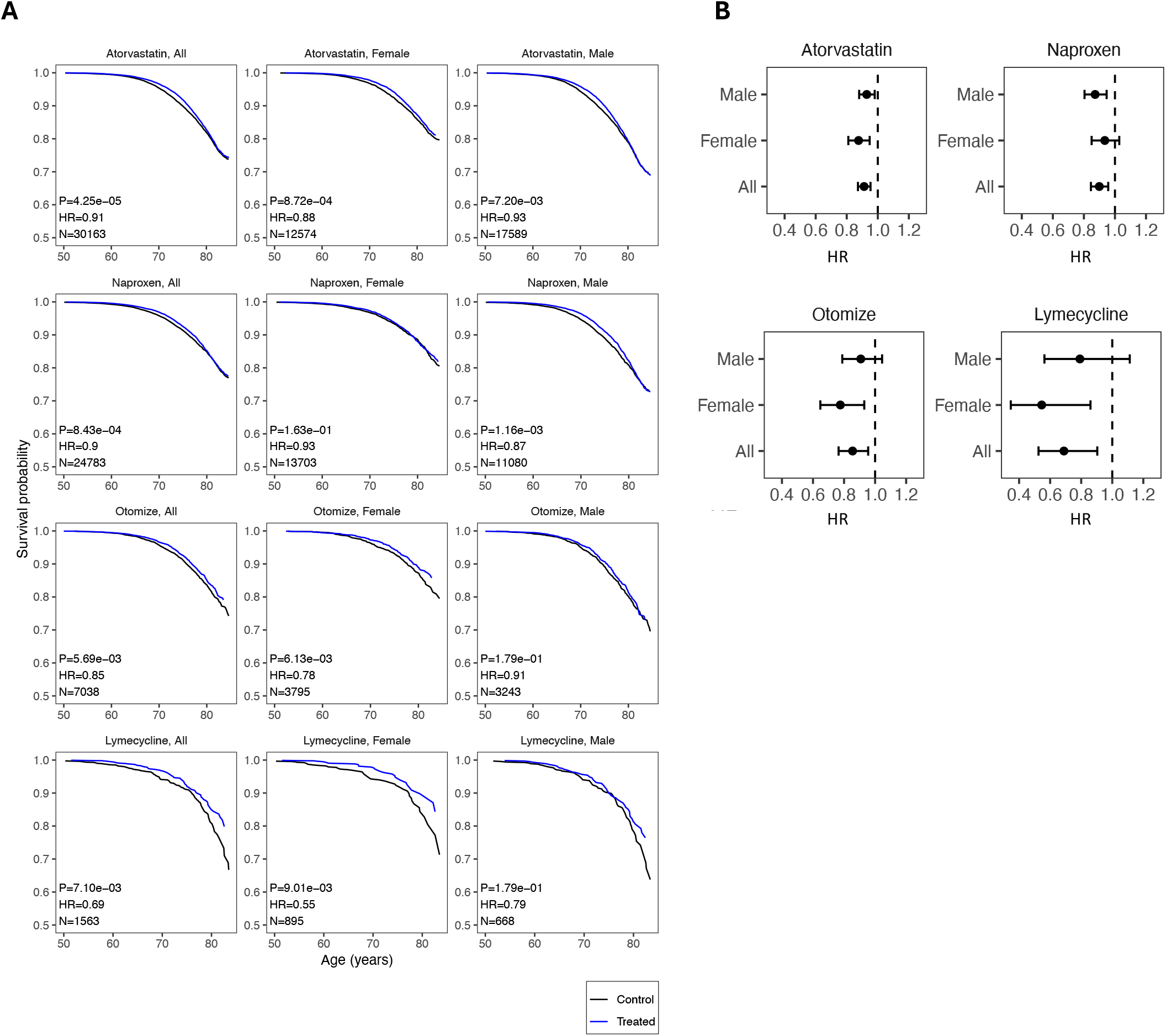
Gender specific effects of prescription drugs associated with decreased mortality. Effect in both (All), female or male only of selected drugs on survival. (**A**) Survival curves. Survival probability vs. Age (years). N: sample size (treated), Hazard-Ratio (HR), > 1 for worse survival, P-value of CoxPH model. Treated group in blue, control in black (**B**) HR with confidence interval 95%. Dotted line HR = 1.

Additionally, we tested the effect of mortality of prescribed drugs, depending on the dosage (**Figure 4, Data Table 3**). For Atorvastatin we observed a J-shaped dose-response effect, with no effect on mortality at 10mg (HR 0.96, CI 0.89-1.02), a reduction in mortality at 20mg (HR 0.87, CI 0.82-0.93), no effect at 40mg (HR 1.03, CI 0.95-1.11) and increased mortality at 80mg (HR 1.17, CI 1.03-1.33). Naproxen reduced mortality to a similar extent at both doses 250mg (HR 0.89, CI 0.80-0.98) and 500mg (HR 0.90, CI 0.84-0.98). Tadalafil reduced mortality at all doses, and most pronounced at 10mg (HR 0.72, CI 0.58-0.89). Sildenafil, likewise, reduced mortality at all doses, with a more pronounced effect at 50mg (HR 0.85, CI 0.75-0.96).

**Figure 4.**
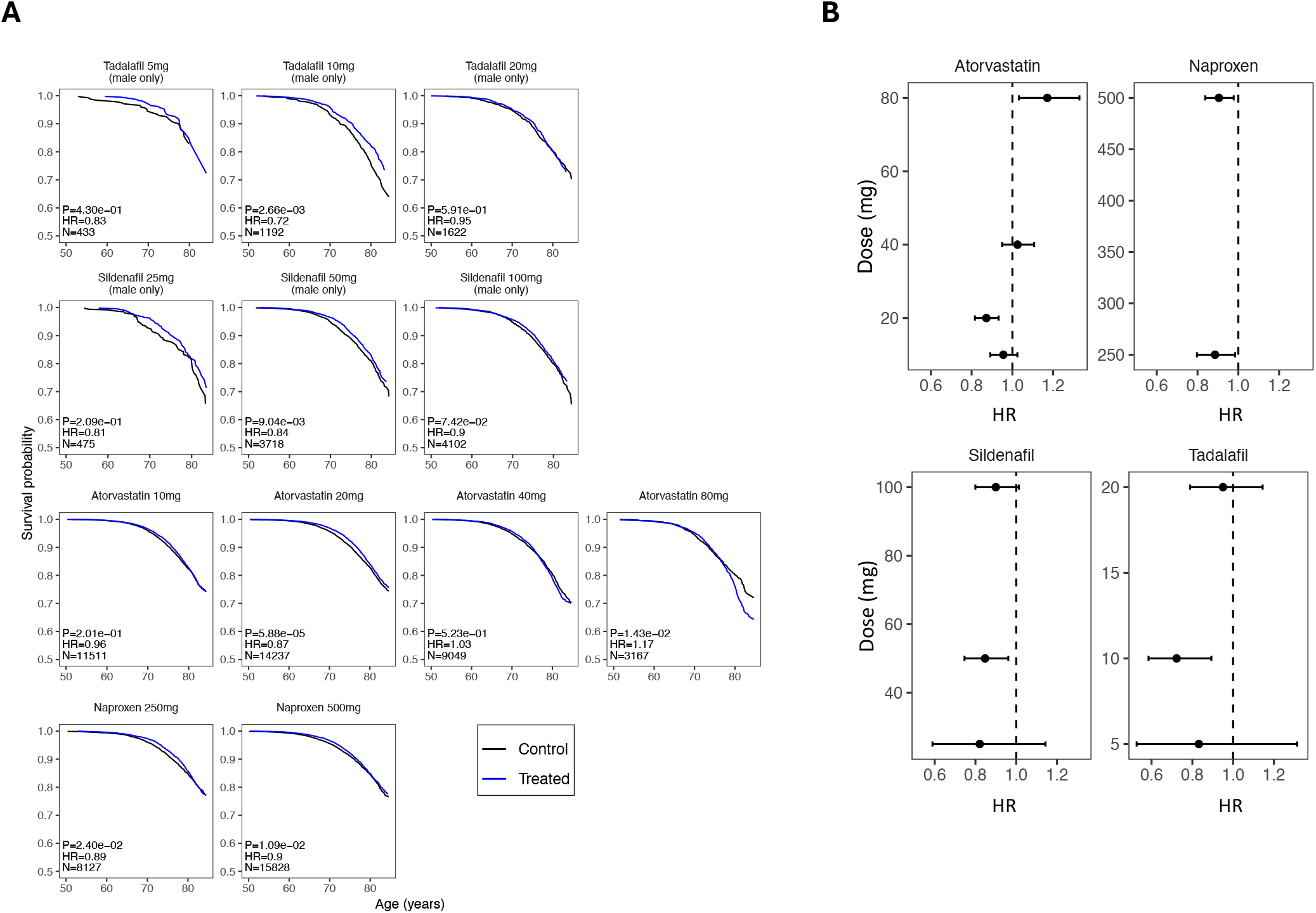
Dose-response effects of prescription drugs associated with decreased mortality. Effect at multiple concentrations (mg) of selected drugs on survival. (**A**) Survival curves. Survival probability vs. Age (years). N: sample size (treated), Hazard-Ratio (HR), > 1 for worse survival, P-value of CoxPH model. Treated group in blue, control in black (**B**) HR with confidence interval 95% in x-axis. Dose (mg) in y-axis. Dotted line HR = 1.

As our findings suggested that there may be a drug class effect on mortality, we performed a pooled analysis, combining drugs from the same class (**Methods**). We selected Statins, PDE5i, Estrogens, given that we observed some positive effect on survival for some of these drugs. We added SGLT2i, and Metformin, due to previous reports of positive effect, and ACEi due to potential reports of positive effects (**Figure 5, Data Table 5**). Statins (HR 0.97, CI 0.94-1.00) and Estrogen (HR 0.76, CI 0.67-0.85), as a class, reduced mortality, so did SGLT2i (HR 0.64, CI 0.45-0.89), although with a much lower sample size. Metformin (HR 1.01, CI 0.95-1.07) had a neutral effect on mortality, while ACEi (HR 1.11, CI 1.06-1.15) was associated with increased mortality.

**Figure 5.**
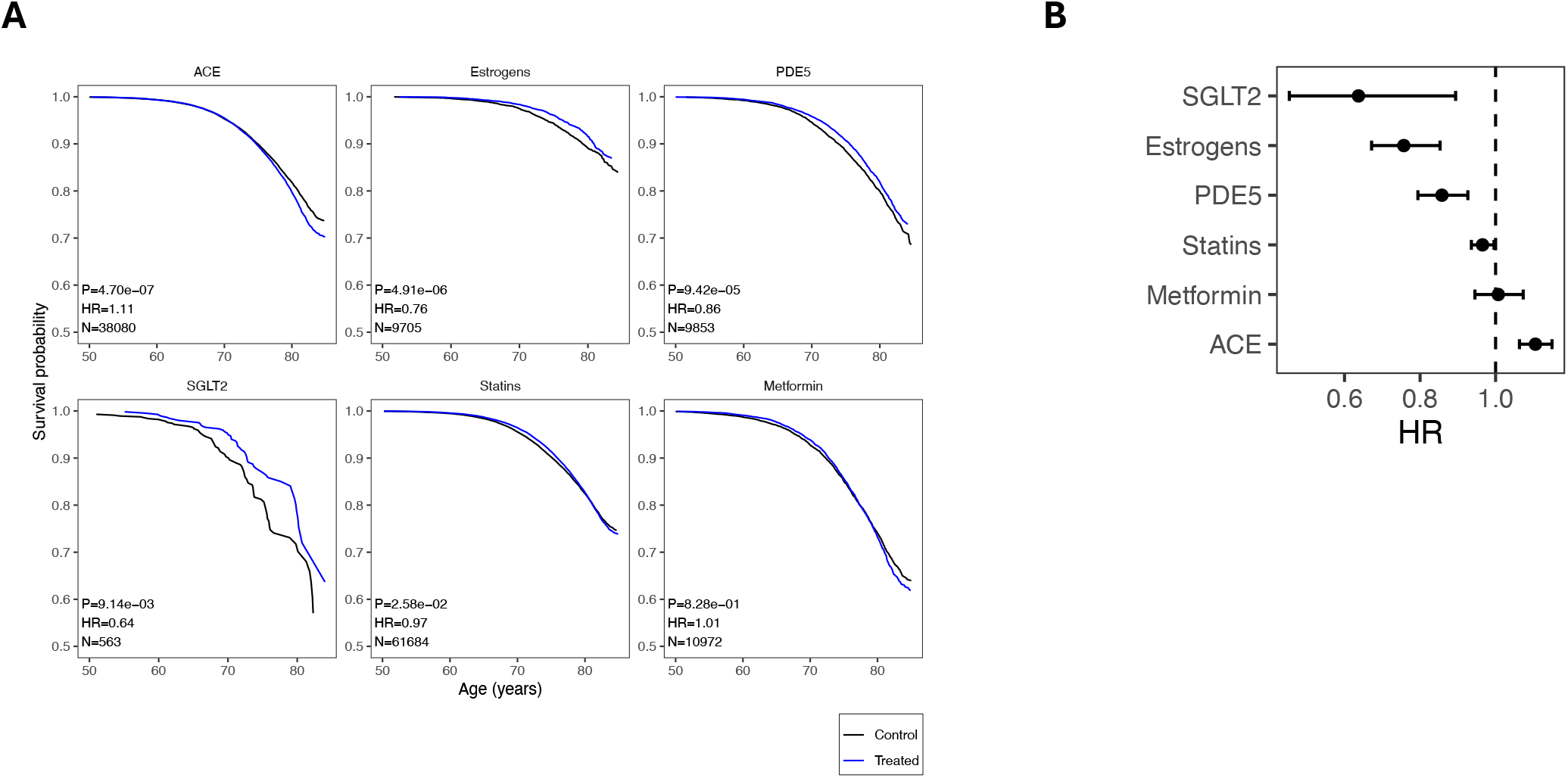
Effect of specific drug classes on mortality. Effect at multiple concentrations (mg) of selected drugs on survival. (**A**) Survival curves. Survival probability vs. Age (years). N: sample size (treated), Hazard-Ratio (HR), > 1 for worse survival, P-value of CoxPH model. Treated group in blue, control in black (**B**) HR with confidence interval 95% in x-axis. Dose (mg) in y-axis. Dotted line HR = 1.

## Discussion

The present study takes advantage of the large UK biobank registry to systematically assess the association between prescribed medications and mortality in the general population. As expected, most prescription drugs are associated with increased mortality. This may in part be driven by the underlying disease the drug is prescribed for, or side effects of the drug. Drugs with increased mortality included opioids (Morphine sulfate, Oramorph), the diuretic Furosemide, pain medication Paracetamol or chronic obstructive pulmonary disease treatment Tiotropium. The detrimental effect of opioids on lifespan has been widely documented^19^. For others, such as widely prescribed Paracetamol, the potential side effect or safety in an older population is still being investigated^20^. In the case of Furosemide, or Tiotropium, the effect might be largely due to the underlying diseases or ailments being treated.

Conversely, we identified 14 drugs associated with decreased mortality in the general population compared to matched controls. These included notably Atorvastatin, Naproxen Sildenafil, and Estradiol. Atorvastatin may work by reducing cardiovascular risk. Indeed, it has been proposed that treating cardiovascular risk factors as early as possible in the general population, with either lipid-lowering medications or anti-hypertensive drugs, may lead to longer lifespans^21^. Yet the benefits of statins on all-cause mortality remains largely debated^14^. PDE5i have also been touted as potential pro-longevity drugs. Notably, in a retrospective study using insurance records, sildenafil was identified to prevent Alzheimer’s disease^22,23^. In another study it was found to prevent CVD and reduce all-cause mortality^24^. Lastly, Estrogen replacement therapy (ERT) have also been reported to have positive effect on mortality in postmenopausal females^25,26^, but there remain active concerns over a potential increased risk of breast cancer^27,28^.

Several drugs had been reported in the past to potentially extend lifespan, compared to non-diseased subjects. In a study from the UK Clinical Practice Research Datalink, metformin-treated diabetic patients had survival rates comparable to (and, among those age > 70, even better than) their matched non-diabetic control group, even though the diabetic patients were more obese and had greater co-morbidities at baseline^17^. Another more recent study found that metformin did not provide survival benefit compared to non-diseased controls, but SGLT2 inhibitors did^11^. In our study Metformin had a neutral effect on lifespan, and SGLT2 inhibitors had a positive impact, but with a lower sample size. Of note the result on metformin has already been questioned in the past^17,18^.

Our study has several limitations. Since most drugs are given in the context of a disease that can limit lifespan, their potential for reducing mortality risk in the general population may be underestimated. Indeed, while it is possible that treating disease-free subjects with the same drugs may increase their lifespan, it was not possible to assess this in the current study and would need prospective randomized clinical studies. Moreover, the results of our study are purely based on retrospective data, and may therefore be confounded by external factors, not recorded in the study. Although we tried to implement an optimal matching strategy, we cannot exclude that for some drugs the matching was not perfect, and therefore control and treated groups did not have the exact same characteristics.

Additionally, despite the large size of the UKBB, for some drugs we were underpowered to detect the effect on mortality as the number of patients receiving the drug was too low. Furthermore, for more recent drug prescriptions, the drug may not have had enough years on the market to be able to influence mortality risk. Moreover, the criteria we used to consider patients taking the drug (more than 3 months between first and last use) allows for drugs being taken intermittently, or for discontinuation of the drug after a 3-month period. Lastly, medication recording in the UKBB was stopped after 2017 and the last few years were missing from our study.

Our study also has several strengths including the large sample size of the UKBB, and the ability the measure the effect on mortality of many drugs at the same time in the same cohort. Another strength was the detailed documentation of key confounding factors, including current smoking, cancer diagnosis, diabetes, gender, age at recruitment, which were used for adjustment in our analyses. Moreover, results of dose-dependent effects, gender-specific effects, and class effect of drugs strengthen the findings of the study.

As the effect of medications on mortality might differ between countries, it would be interesting to validate these results by performing similar analyses in health registries from other countries. As certain biological mechanisms can improve health early in life but impair it later^29,30^, it may be interesting to also investigate age dynamics of the observed associations. Drug interactions could also be explored as part of future studies but may be hampered by low sample size. Similarly, associations between drug prescription and particular cause of death could be explored. Lastly the benefit on lifespan observed for several drugs in this retrospective study could only be truly determined by performing randomized controlled trials (RCT). Such studies would aim at treating a healthy aging population with a drug during a sufficiently long follow-up to be able to observe the effect on all-cause mortality. The Targeting Aging with Metformin (TAME) trial^31^ has been proposed for many years based on such concept, but to our knowledge this trial has not started yet, nor has any other trial of this kind.

## Methods

### Dataset

The data from UKBB was retrieved on October 24, 2023. The fields selected from the participant table were Participant ID, Date of attending assessment center, Age at recruitment, Year of birth, Month of birth, Age at Death, Date of Death, Underlying primary cause of death, Sex, Genetic principal components 1-10, Summed MET minutes per week for all activity, Body mass index BMI, Townsend deprivation index at recruitment, Pack years of smoking, Smoking status, Sleep duration, Cancer diagnosed by doctor, Fractured broken bones in last 5 years, Other serious medical condition / disability diagnosed by doctor, Vascular / heart problem diagnosed by doctor, Blood clot / DVT / bronchitis / emphysema / asthma / rhinitis / eczema / allergy diagnosed by doctor, Diabetes diagnosed by doctor, Age diabetes diagnosed, Alcohol intake frequency, Diagnoses – ICD10. The prescription data was obtained from the Primary Care Linked Data and the fields selected from the prescription data were Participant ID, Date prescription was issued and Drug name.

### Inclusion/Exclusion of covariates

For these fields to be used in the covariate analysis, they were to present one of two shapes: binary and quantitative covariate. All the fields representing comorbidities at recruitment were transformed into binary covariates (is affected or not affected by this comorbidity). All the fields with quantitative values were left as is. Finally, more complex fields like smoking status were transformed into multiple binary covariates (current smoker and never smoked) and alcohol intake frequency was transformed into monthly alcohol intake, following the mean value of the corresponding code.

Out of all of these, the fields with the most missing data were Pack years of smoking and Summed MET minutes per week for all activity. A univariate survival analysis showed that these covariates had a very low impact on the survival rate, and they were excluded.

### Inclusion/Exclusion of participants

We censored every participant entry where one of the covariates was missing, as described in the covariate analysis section. Every participant whose death was self-inflicted or the result of an accident (ICD10 codes starting with O, Q, S, T, V, W, X, Y) were also censored. From the 501’169 starting participants of the UK biobank, we were left with 480’444.

### Covariate analysis

The covariates selected for the analysis were Current smoker, Cancer diagnosed, Age at recruitment, Diabetes diagnosed, Sex (is male), Other serious medical condition / disability, Vascular / heart problems diagnosed, Never smoked, Townsend deprivation index, Fractured broken bones in last 5 years, Blood clot / DVT / bronchitis / emphysema / asthma / rhinitis / eczema / allergy diagnosed, Body mass index BMI, Alcohol intake monthly frequency, Genetic principal components 1-10, Sleep duration.

We conducted a Cox Proportional Hazards (CoxPH) multivariate survival analysis and classified the covariates under three categories depending on their effect on the survival rate. With a hazard ratio greater than 1.5, covariates “Current smoker”, “Cancer diagnosed”, “Age at recruitment”, “Diabetes diagnosed”, and Sex (is male) were called high-priority covariates. With a hazard ratio greater than 1.1, covariates “Current smoker”, “Cancer diagnosed”, “Age at recruitment”, “Diabetes diagnosed”, and Sex (is male) were called high-priority covariates Other serious medical condition / disability, “Vascular / heart problems diagnosed”, “Never smoked”, “Townsend deprivation index”, “Fractured broken bones in last 5 years”, “Blood clot / DVT / bronchitis / emphysema / asthma / rhinitis / eczema / allergy diagnosed” were called intermediate-priority covariates. All the others were called low-priority covariates.

### Matching

Subjects that were prescribed the drug of interest for at least 3 consecutive months were individually matched with subjects that were not prescribed that drug (or prescribed for less than 3 months) using an algorithm called Nearest Neighbor Covariate Matching (NNCM)^32–34^ . Given how important the different covariates are on the survival rate of individuals, this matching attempts to make sure that the individuals with prescription are compared to individuals presenting a similar subset of covariates.

The covariates were assigned different levels of priority (defined in the covariate analysis), relative to their impact on the survival rate of individuals. The distance metric used in the matching algorithm was adapted to give more weight to the covariates with a higher priority. In other words, the higher the priority on a covariate, the stricter the matching will be relative to that covariate.

Example of this matching is presented in the appendix (**Figure S2**). The difference in matching between higher priority covariates and lower priority ones can clearly been seen. To achieve an optimal matching strategy, we decided to include all the participants in the cohort, includes ones for which no prescription data was available. We cannot exclude that some of the matched participants were also being prescribed the drug in question.

### Drug name curation

A challenge encountered with the drug prescription table is that the same drug can be presented under several different names. An example of this with Omeprazole was the presence of at least 20 variants of “Omeprazole 20mg capsules”, such as “Omeprazole Cap 20mg” or “Omeprazole Multiple Unit Pellet System Dispersible Tablets 20mg”.

To circumvent this issue, two sets of some techniques were used. First automatic techniques such as capitalization and the removal of special characters. Then finally, the remaining drug names were curated matched ‘by hand’. This method allowed to not only study the effect of drugs in general but also to study the effect of the drugs by concentration.

We limited our analysis to the top 1000 most used drugs in the raw prescribed data after the first automatic round of curation. The second round of handmade name curation brought that number down to 543 unique drugs and 829 unique drugs-concentration couples. Finally, it was decided to only keep the drugs that were prescribed to at least 500 patients for at least 3 months, which gave the final number of 406 drugs.

### Survival analysis

The survival analysis was obtained by comparing two groups for each drug. The first group is comprised of individuals that have been using the drug of interest for at least three months. The second group is matched individuals that haven’t been using the drug of interest. The results of the survival analysis were obtained by conducting a univariate CoxPH analysis on these two groups.

We conducted three different analyses. The first was on the drug use regardless of concentration, the second one was on the drug and their specific concentration, and the last one was on specific drug groups. For each of these, we analyzed the results on the whole prescription group as well as on the male patients only and female patients only.

The specific drug groups for the third analysis are statins, with Atorvastatin, Fluvastatin, Pravastatin, Rosuvastatin and Simvastatin; SGLT2 inhibitors (Canagliflozin, Empagliflozin, Dapagliflozin); PDE5 inhibitors, with Tadalafil, Sildenafil and Vardenafil; ACE inhibitors, with Captopril, Enalapril, Fosinopril, Lisinopril, Perindopril, Ramipril and Trandolapril; Metformin; Estrogens, with Estraderm, Estradiol, Estriol and Vagifem.

### Multiple comparison correction

Assessing significance of results on a multiple comparisons problem can be challenging since having a low threshold for the p-value might not be enough to dodge type I errors^35^ For these survival analyses, we decided to implement the Benjamini–Hochberg procedure^36^ with a control level alpha equal to 0.05.

**Figure S1.**
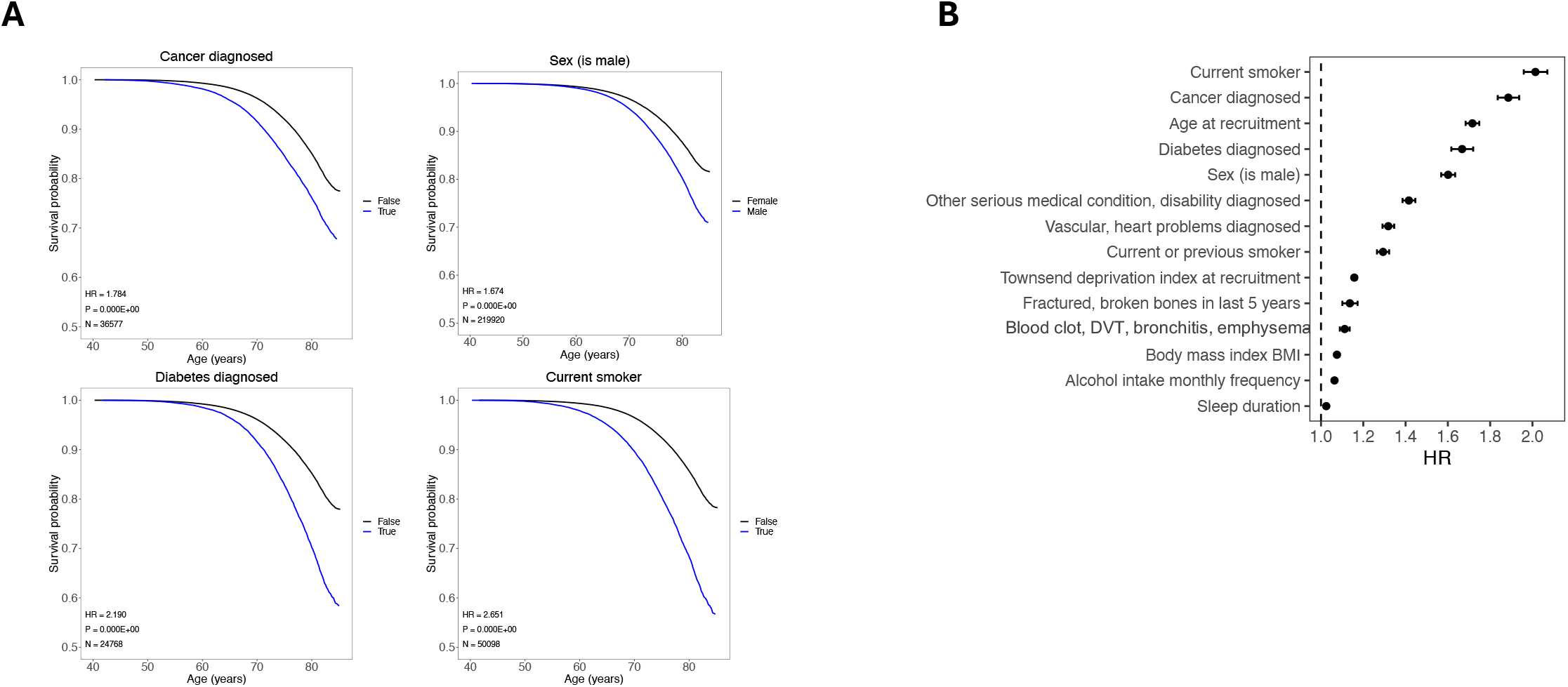
Effect of main covariates on mortality. Effect if selected covariates on survival. (**A**) Survival curves. Survival probability vs. Age (years). N: sample size (treated), Hazard-Ratio (HR), > 1 for worse survival, P-value of CoxPH model. Treated group in blue, control in black (**B**) HR with confidence interval 95% in x-axis for top covariates. Dotted line HR = 1.

**Figure S2.**
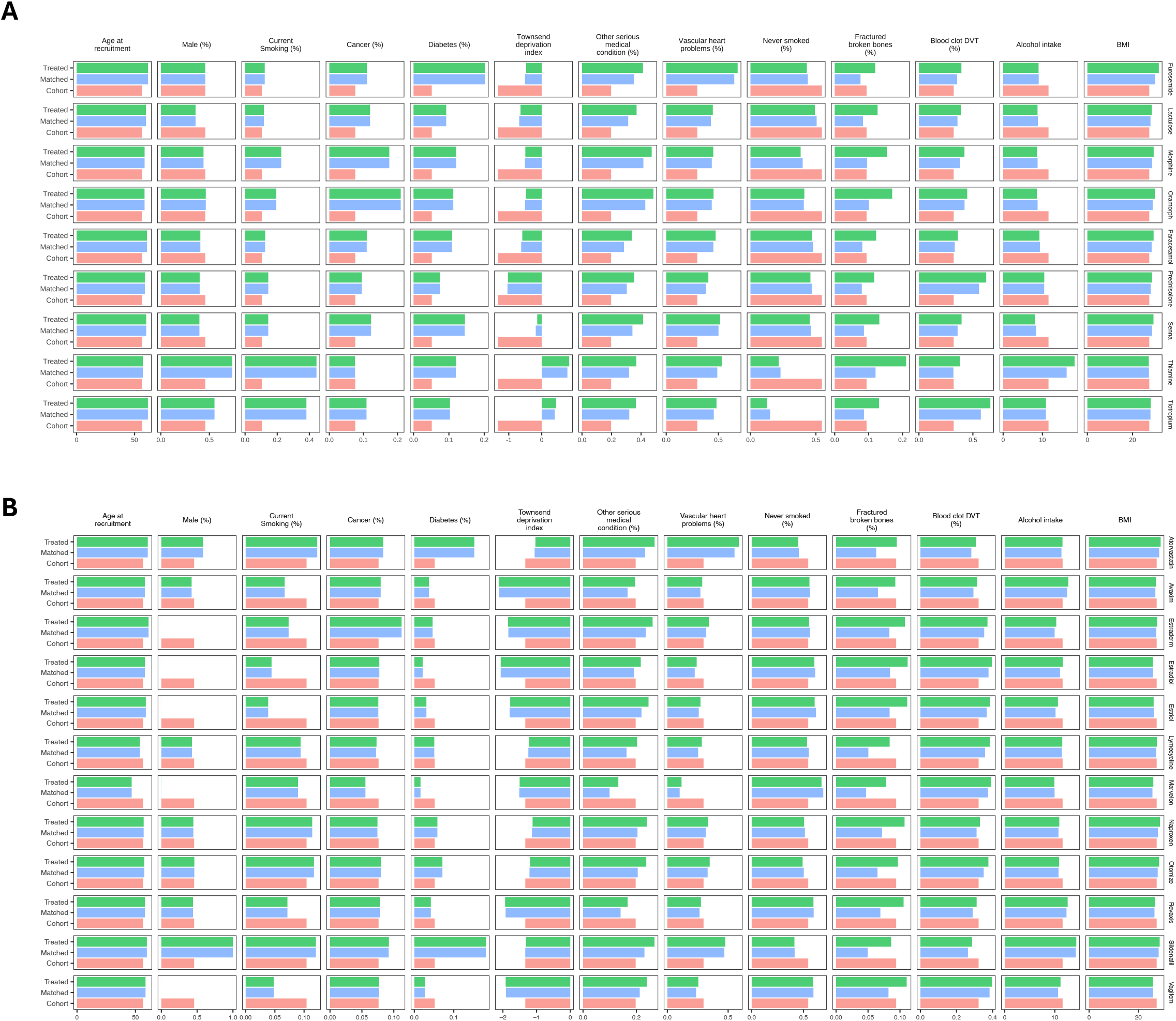
NNCM matching results. (**A**) Matching of prescription drugs associated with decreased lifespan. (**B**) Matching of prescription drugs associated with increased lifespan. Whole cohort in red, treated group in green, matched control group in blue.

**Table S1.**
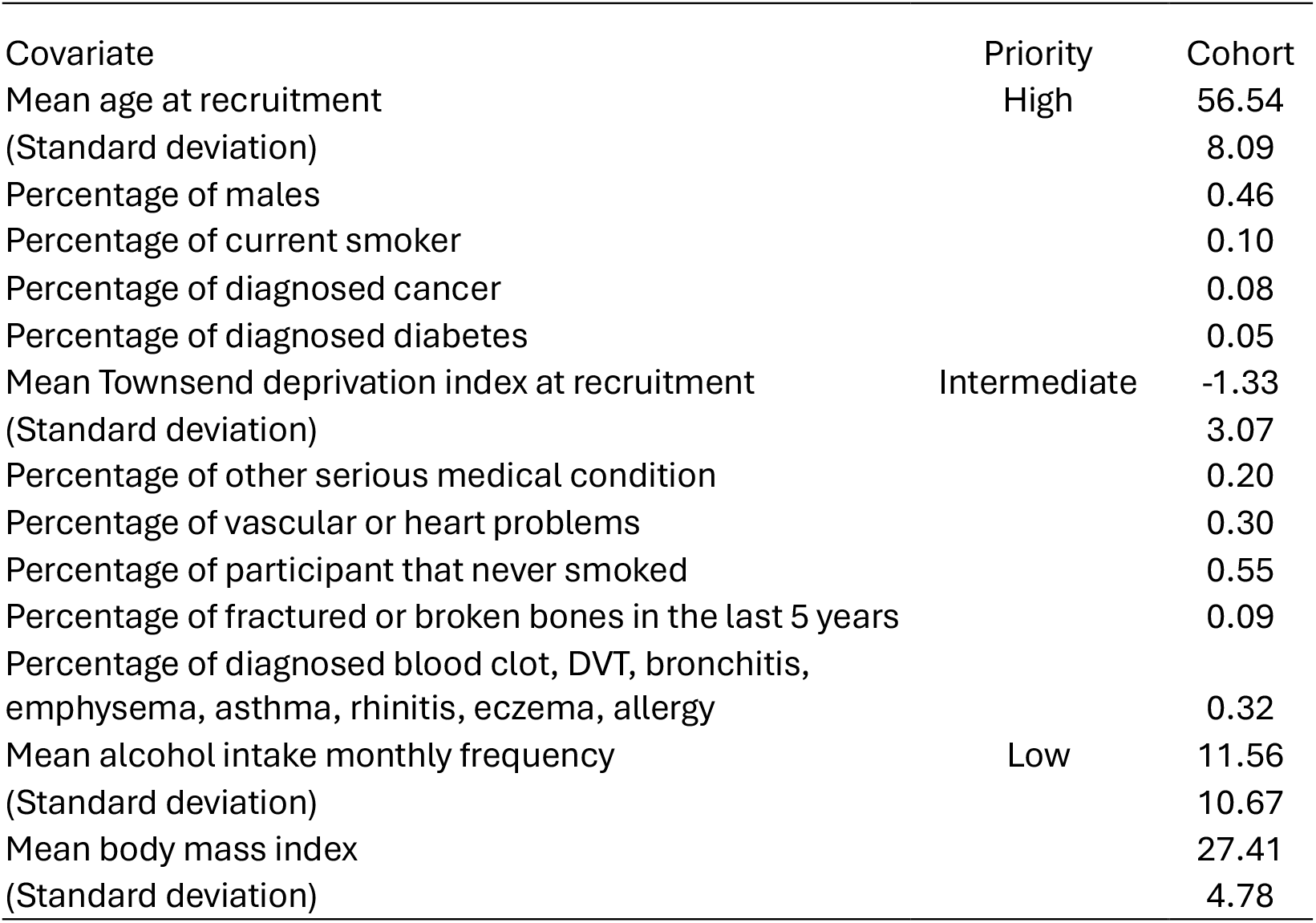
Cohort characteristics. Distribution of main covariates in the whole cohort, sorted by priority.

## Data Availability

All data produced in the present work are contained in the manuscript. No additional data available.

## Acknowledgements

We thank the participants and data curators of the UK Biobank. We thank all the team members of EPITERNA.

## Conflict of Interest

UK Biobank was established by the Wellcome Trust medical charity, Medical Research Council, Department of Health, Scottish government, and Northwest Regional Development Agency. EPITERNA is a private for-profit entity, but with no outstanding financial interest in the presented prescription drugs.

### Author contribution

J. M. performed data and statistical analysis, visualization and co-wrote the manuscript. Y.R. and H.B.F critically revised the manuscript. A. O. co-supervised the work, designed the study, and co-wrote the manuscript. K. P. co-supervised the work, designed the study, performed data analysis, and co-wrote the manuscript.

### Ethics approval

UK Biobank has obtained ethics approval from the Northwest Multi-Centre Research Ethics Committee (approval number: 11/NW/0382) and has obtained informed consent from all participants.

### Data access

Data access was granted as “Approved Research” study from the UK biobank, principal investigator: Dr Kevin Perez (EPITERNA SA), Approved Research ID: 107065, Approval date: August 17th, 2023.

### Funding

UK Biobank was established by the Wellcome Trust, Medical Research Council, Department of Health, Scottish government, and Northwest Regional Development Agency. It has also had funding from the Welsh assembly government and the British Heart Foundation. The data access and analyses in this study were funded by EPITERNA.

